# Saliva direct PCR protocol for HLA-DQB1*02 genotyping

**DOI:** 10.1101/2023.10.25.23297535

**Authors:** Angeles Carrillo, María Jimena Manzur, Maximiliano Juri Ayub

**Affiliations:** Laboratorio de Desarrollo de Diagnósticos Moleculares, Área Biología Molecular, Facultad de Química, Bioquímica y Farmacia, Universidad Nacional de San Luis. Ejército de los Andes 950, D5700HHW San Luis, Argentina

## Abstract

Celiac Disease (CD) is an immune disorder that is triggered by gluten ingestion in genetically predisposed individuals. The HLA-DQB1*02 allele is the main predisposing genetic factor, and a candidate for first-line genotyping screening. We designed and validated a simple, DNA purification-free PCR protocol directly from crude saliva, enabling the detection of the DQB1*02 allele. This assay also distinguishes homozygous from heterozygous carriers. We propose this method for use in mass screening and/or epidemiological studies.

## INTRODUCTION

Celiac Disease (CD) is an immune disorder triggered by gluten ingestion in genetically predisposed individuals (1). Four alleles at two HLA *loci* (DQA1*03 and *05, DQB1*02 and 03:02) are associated with CD. Moreover, the absence of these alleles has a high negative predictive value. HLA-DQB1*02 is by far the strongest predictor of genetic susceptibility, being present in 90-95% of patients with CD (2). Therefore, it has been suggested that massive genetic testing for this allele would be desirable if cost-effective strategies are available (2-6). Moreover, a two-step screening strategy has been proposed for children: a first-line screening for HLA-DQB1*02, followed by prospective serological studies only for DQB1*02 carrying individuals (7).

In the present study, we describe a simple, fast and affordable saliva direct qPCR protocol for HLA-DQB1*02 detection, exploiting a 52 bp insertion located in the second intron of this allele. This simple and low-cost protocol can be used for first-line clinical and epidemiological analyses.

## METHODS

### Saliva collection

Saliva was self-sampled by volunteers with informed consent. Smoking, eating, and drinking were avoided for 1 h prior to collection. Saliva was allowed to naturally accumulate in the mouth and was self-collected in sterile, RNase and DNase-free 1.5 ml plastic tubes.

### DNA purification from saliva

Total nucleic acids were purified from 140 μl of saliva using a standard column purification kit, according to the manufacturer’s instructions (http://www.pb-l.com.ar/productos/puro-virus-rna).

### Sequence genotyping HLA-DQB1 alleles

Purified DNA was used as a template for amplifying a 254 bp region of HLA-DQB1 using the following primers: <u>**T7prom**</u>-DQB1-Fw: <u>**TAATACGACTCACTATAGGG**</u>GATTCCYCGCAGAGGATTTCG (500 nM), DQB1-Rev1: CCAACTGGTAGTTGTGTCTGC (250 nM) and DQB1-Rev2: CCACCTCGTAGTTGTGTCTGC (250 nM). Cycling conditions were 3 min at 95ºC and 40 cycles of 95ºC for 10s/63 ºC for 30s, using a LightCycler Multiplex RNA Virus Master (Roche). Amplicons were subjected to Sanger sequencing using the T7promoter primer at the Macrogen facility. By analyzing the polymorphic sites in the chromatograms, both alleles in each sample were resolved.

### Design of HLA-DQB1-specific PCR

The HLA-DQB1 alleles were retrieved from the IPD-IMGT/HLA Database (https://www.ebi.ac.uk/ipd/imgt/hla/alleles/). Sequences were aligned using MAFFT (https://www.ebi.ac.uk/Tools/msa/mafft). Sequence alignment was visualized using MEGA 6 software (8), revealing a DQB1*02-specific 52 bp insertion which would be adequate for distinguishing this allele from others by PCR (**Figure 1**). Therefore, we designed two PCR primers (DQB1*02-Fw: CATCAGAGATTAGCATCACCAC and DQB1*02-Rev: CTGATTACTTAAAGTGTTTATCATGTAC) flanking this insertion, which should amplify a fragment of 121 bp (for DQB1*02) or 69 bp (for other alleles). Using *in silico* prediction of Tm (https://www.dna-utah.org/umelt/quartz/um.php), the DQB1*02 amplicon (121 bp) was expected to exhibit a Tm approximately 5ºC higher that of other alleles (69 bp).

**Figure 1.**
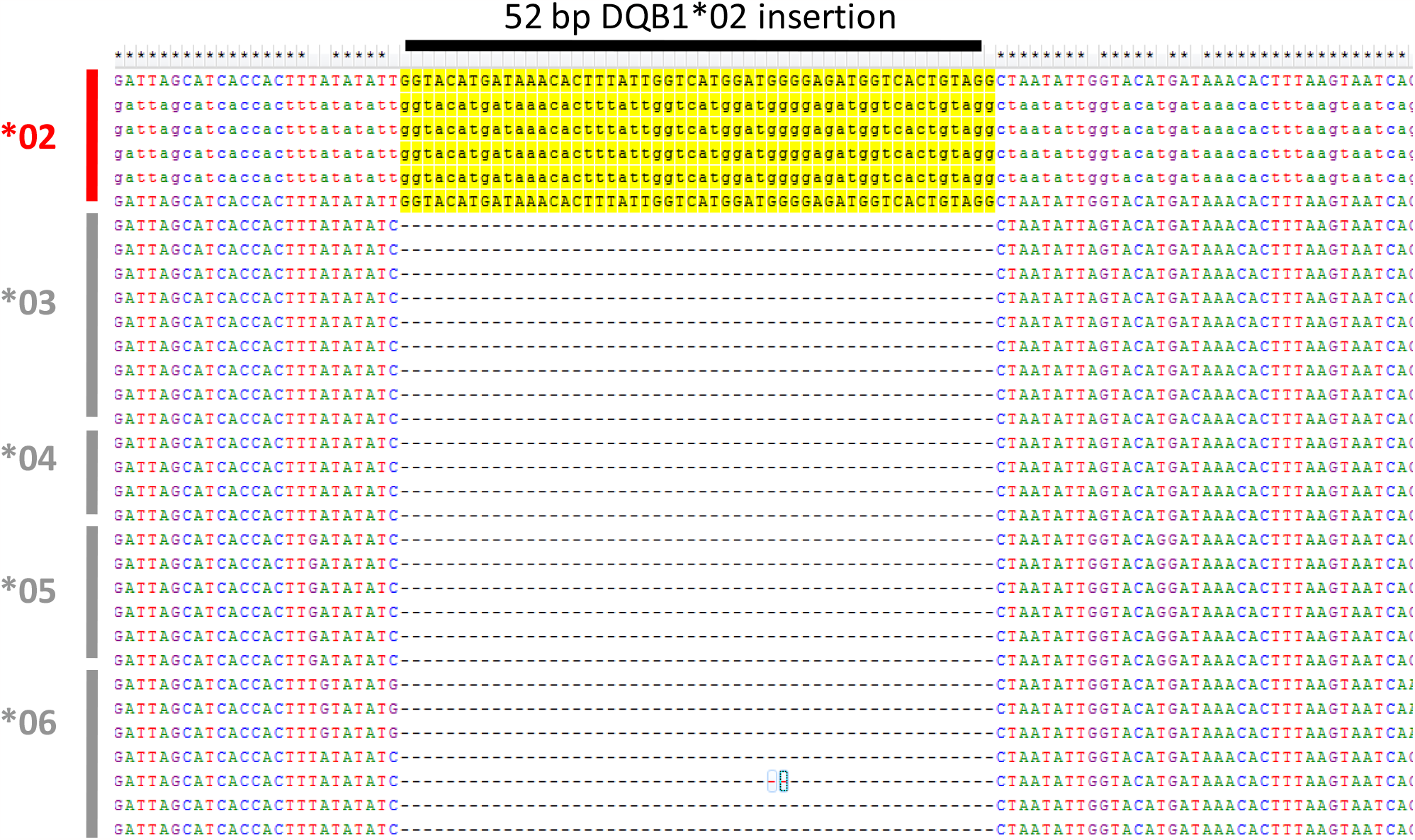
Sequence alignment of DQB1 alleles showing the 52 bp insertion unique to DQB1*02.

### Sequence genotyping HLA-DQB1 alleles

Primers DQB1*02-Fw and DQB1*02-Rev (500 nM each) were used for PCR amplification in 20 ul reactions in the presence of 1 μl EvaGreen under the following cycling conditions: 1 min at 95ºC and 40 cycles of 95ºC for 10s/60 ºC for 30s. DNA purified from saliva as described previously or, alternatively, crude saliva was used as template. After amplification, a melting step from 60 ºC to 90 ºC was performed, and fluorescence was recorded every 0.5 ºC. Derivatives of the melting curves were plotted to discriminate amplicons derived from DQB1*02 from other alleles. Two commercial PCR master mixes yielded similar positive results: the LightCycler Multiplex RNA Virus Master (Roche) and qScript XLT 1-Step RT-qPCR ToughMix (QuantaBio).

## RESULTS

Saliva from different individuals was genotyped by PCR amplification and sequencing, as described in the Methods section. Next, samples harboring (in homozygosity or heterozygosity) or lacking the DQB1*02 allele (as determined by Sanger sequencing) were selected and analyzed by DQB1*02 genotyping PCR. **Figure 2** shows the representative melting curves obtained from homozygous (*02-02) or heterozygous (*02-X) positive and negative (*X-X) samples. Non-DQB1*02 alleles showed a melting peak around 70 ºC whereas the DQB1*02 allele was clearly detected as a distinct 75 ºC peak. As expected, both peaks were observed in heterozygous. Interestingly, using 1ul of crude saliva yielded identical results to using purified DNA.

**Figure 2.**
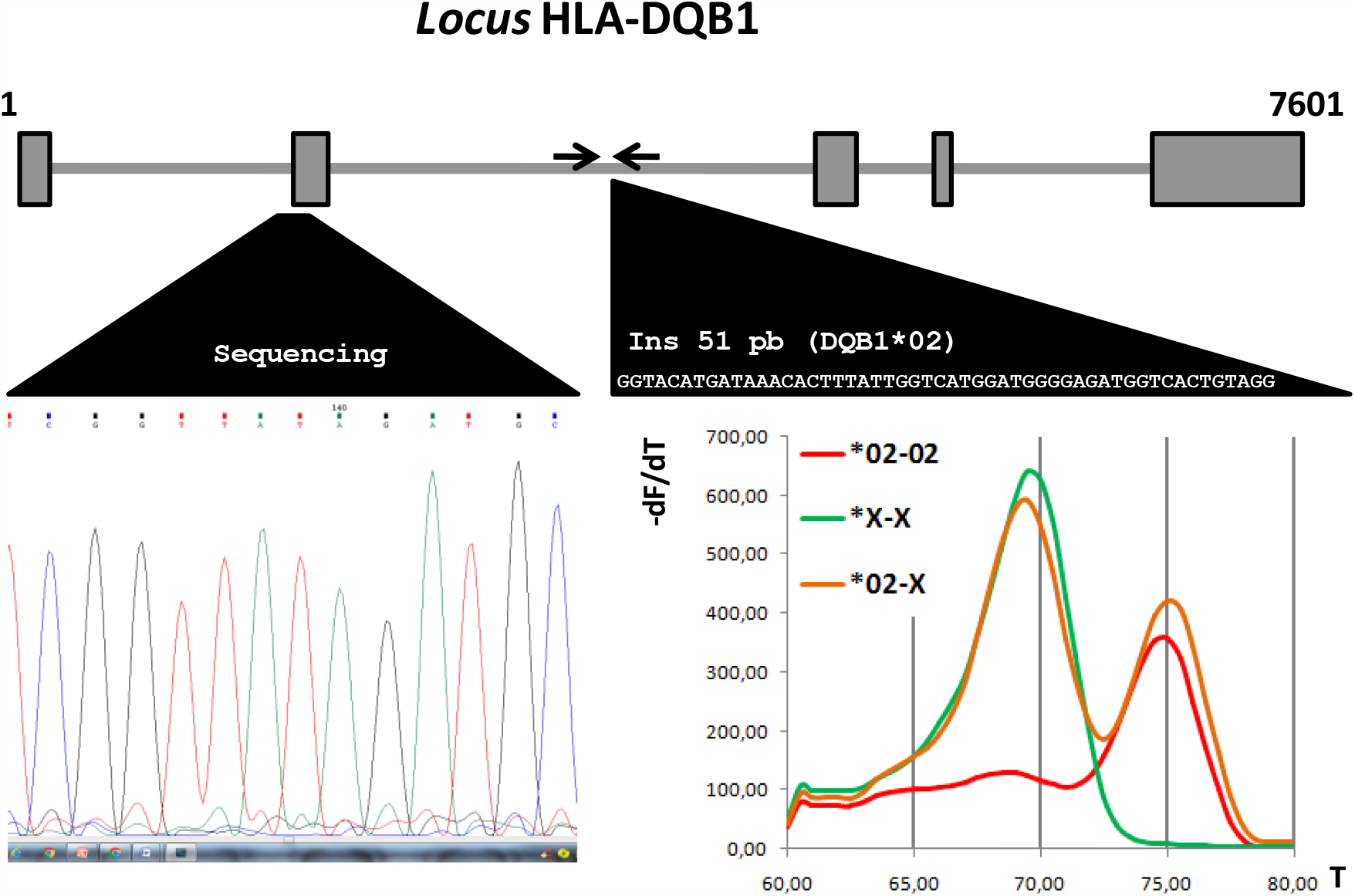
Schematic representation of the exón-intron structure of HLA-DQB1 gene and genotyping strategies. Genomic (NT_167248) and mRNA (NM_001243962) sequences were analyzed using SPLIGN (https://www.ncbi.nlm.nih.gov/sutils/splign/splign.cgi) to identify exonic (grey boxes) and intronic (line) regions. The amplicons used for Sanger sequencing (left) and melting genotyping (right) are depicted. Melting peaks of representative samples are shown.

**Figure 3.**
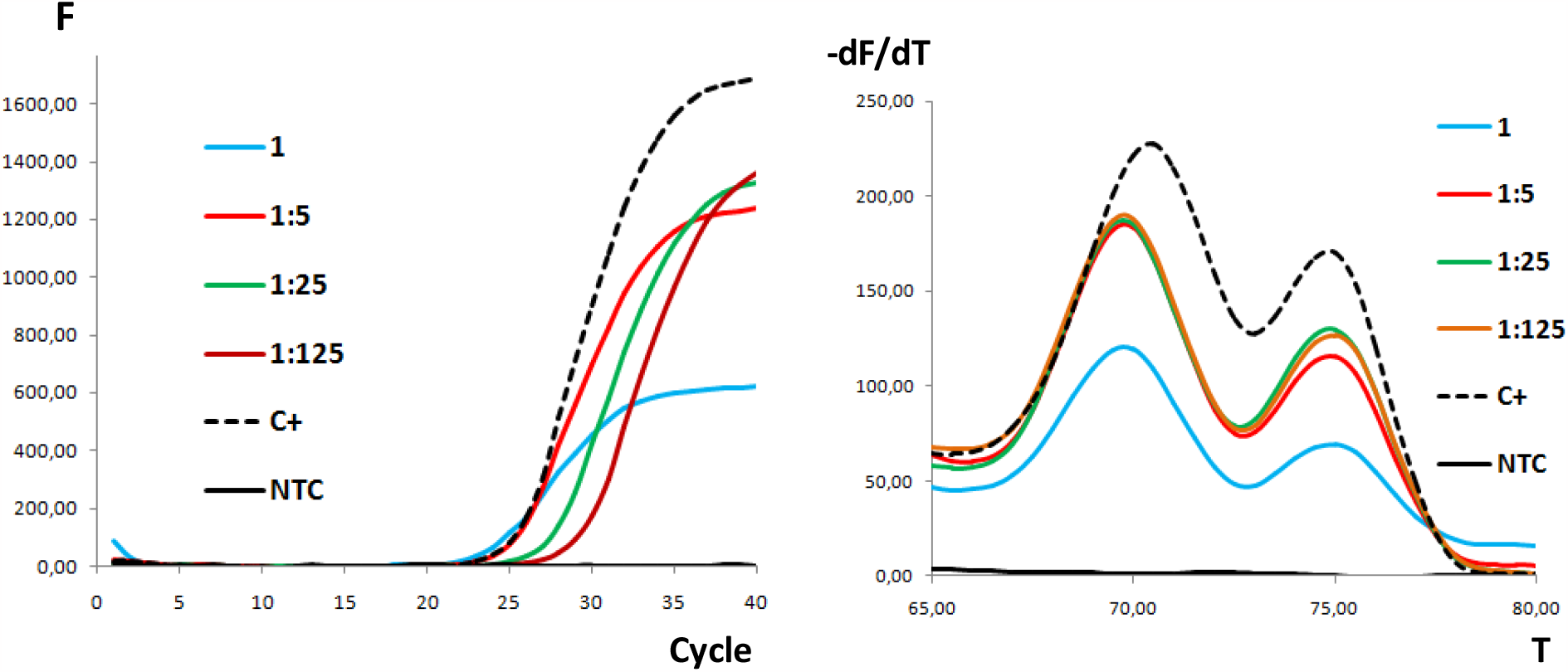
Five microliters of pure saliva (1) and 1:5 serial dilutions were used as templates in 20 μl reactions, as described previously. Water (NTC) and purified DNA (C+) were used as negative and positive controls, respectively. The amplification (left) and melting curves (right) are shown. All the samples yielded significant signals.

In summary, our HLA-DQB1*02 genotyping assay has the following advantages for general screening and epidemiological studies:

1. It requires a minimal amount of saliva, which can be easily provided by patient without assistance from professionals.
2. PCR can be performed using crude saliva, without any processing or purification steps, saving cost and work time, and minimizing the risk of cross-contamination among samples.
3. No control reaction (*e*.*g*. globin or actin gene) is required, because amplification should be detected for any genotype, ruling out the possibility of false negative results.
4. Homozygous and heterozygous DQB1*02 carriers are clearly distinguishable.
5. The protocol was highly tolerant to changes in the amount of template used, yielding clear results for both pure and highly diluted saliva samples.

Finally, the general strategy of exploiting specific InDels for detecting specific alleles from a locus can be extended to other disease-associated genes.

## Data Availability

All data produced in the present work are contained in the manuscript

## ACKNOWLEDGEMENTS

We are deeply indebted to the volunteers who participated in the study. A.C. is a laboratory technician at the Universidad Nacional de San Luis (UNSL). M.J.M. and M.J.A. are members of the Consejo Nacional de Investigaciones Científicas y Técnicas (CONICET) career. Specific funding for this project provided by the UNSL is also acknowledged.

